# A prototype vaccination model for endemic Covid-19 under waning immunity and imperfect vaccine take-up

**DOI:** 10.1101/2021.11.06.21266002

**Authors:** John Dagpunar, Chenchen Wu

## Abstract

In this paper, for an infectious disease such as Covid-19, we present a SIR model which examines the impact of waning immunity, vaccination rates, vaccine efficacy, and the proportion of the susceptible population who aspire to be vaccinated. Under an assumed constant control reproduction number, we provide simple conditions for the disease to be eliminated, and conversely for it to exhibit the more likely endemic behaviour. With regard to Covid-19, it is shown that if the control reproduction number is set to the basic reproduction number (say 6) of the dominant delta (B1.617.2) variant, vaccination alone, even under the most optimistic of assumptions about vaccine efficacy and high vaccine coverage, is very unlikely to lead to elimination of the disease. The model is not intended to be predictive but more an aid to understanding the relative importance of various biological and control parameters. For example, from a long-term perspective, it may be found that in the UK, through changes in societal behaviour (such as mask use, ventilation, and level of homeworking), without formal government interventions such as on-off lockdowns, the control reproduction number can still be maintained at a level significantly below the basic reproduction number. Even so, our simulations show that endemic behaviour ensues. The model obtains equilibrium values of the state variables such as the infection prevalence and mortality rate under various scenarios.

## 1 Objectives, methodology, and main findings

In this paper, we develop a vaccination model that examines the impact of different controls on the long-term endemic behaviour of Covid-19. These controls include different levels of vaccine waning rates, vaccine efficacy, rate of vaccination, target vaccination coverage, and control reproduction number. As many have observed, there is still uncertainty about efficacy and waning. While reasonable estimates are available for efficacy of a given vaccine against symptomatic disease, there is less confidence in estimates against infection, hospitalisation, and death, and these contribute to uncertainty in prediction, even in the short term. Therefore, it is stated at the outset, this is not a predictive model, more one to aid understanding. Various scenarios are illustrated for the UK situation, although the model is not confined to any geographical region.

The primary interest is in a long-term behaviour. It is generally believed that Covid-19 will be around for a long time, so policymakers need to decide on a level of long-term controls that will deliver a defined balance between prevalence of the disease and economic and other damage resulting from such controls. The original wild version of the virus had a reproduction number of around 3 (Billah, Miah, and Khan, 2020). Theoretically, it might have been possible to eliminate the disease in the long term with vaccination alone. Now, with the delta variant having an estimated basic reproduction number of between 5 and 8 (Liu and Rocklöv, 2021), disease elimination seems extremely unlikely, unless one were prepared to live under extreme controls in the long term.

Using the model, we illustrate circumstances in which long honeymoon periods of low prevalence can exist between waves of infection whose successive peaks decline. Such model behaviour has also been observed for childhood diseases such as measles and mumps, under an assumption that immunity from vaccination may not be lifelong; see, for example, Scherer and McLean (2002). We show that increasing target vaccine coverage from 65% to 95% might reduce long-term death rates by a factor of 5, while when target vaccination coverage is 65%, equilibrium death rates can change by factors of 14 and 3 depending upon assumptions made about waning and vaccination rates respectively. Different vaccines have different efficacies against infection and so the finding that changing that from 65% to 90% in one modelled scenario, can change the equilibrium death rate by a factor of 10, might be seen as having important implications for distribution of limited supplies of different vaccines having different efficacies, amongst different risk groups. Assuming a basic reproduction number of around 6, we examine a hypothetical scenario where a control reproduction number less than that is sustainable in the long term. According to one modelled scenario, changing that from 4 to 3 might reduce the equilibrium death rates by a factor of 4. Finally, the model allows one to compare equilibrium death rates under vaccination with those under no vaccination. In a base case scenario, adjusted to a vaccination coverage of 95%, the ratio was 29.

The structure of the paper is as follows. In section 2 we summarise the relevant literature in this area. In section 3, the mathematical model is developed, with results shown in section 4. In section 5, we have further discussion related to the disease-free equilibrium, death rate, and limitations of our model. Section 6 summarises and concludes.

## 2 Background and literature on vaccine models

There is an established history of using mathematical models to study the evolution of epidemics and the effectiveness of vaccination programmes in epidemic control. In searching the literature, we were particularly interested in identifying those studies that included waning of immunity, which seems to be an important issue with Covid-19; see, for example, Altmann and Boyton (2021).

Scherer and McLean (2002) developed SIR vaccination models to answer basic questions regarding diseases such as measles, mumps, and rubella, where a certain proportion are vaccinated just once in early childhood, and where an assumption of lifelong immunity is relaxed by introducing waning of immunity. They show how to calculate thresholds for disease-free and endemic behaviour. If the threshold is only just exceeded, then that typically results in long honeymoon periods of low prevalence between successive subsequent waves. Feng, Towers, and Yang (2011) develop SIR and stochastic simulation models for pandemic influenza with seasonal transmission rates, vaccination, and antiviral treatments included. As waning immunity is not modelled, the question of long-term endemic levels of disease is not considered.

When we focus on literature for Covid-19 and related vaccination programmes, SIR and SEIR models are frequently used. Hollingsworth, Okamoto, and Lloyd (2020), in a non-immunising environment, examine the combined effect of transient controls and waning immunity, that can result in accumulation of susceptibles and a resultant emergence of larger than expected waves, the so-called divorce effect. Annas et al. (2020) and Batistela et al. (2021) both model vaccination by moving people from a susceptible compartment to a recovered one, which implicitly assumes 100% vaccine efficacy and both include theoretical stability analysis of disease-free and endemic equilibrium. Giordano et al. (2021) use a compartmental vaccine model to predict outcomes in Italy from initial roll-out from April 2021 until January 2022, under various Non-Pharmaceutical Interventions (NPIs) control strategies and rates of vaccination of the population. Their results confirm the importance of not releasing NPIs until a sufficient proportion of the population has been vaccinated. The methodology optimistically assumes 100% vaccine efficacy and no waning of immunity after vaccination or infection.

Models developed by Xu, Wu, and Topcu (2021), Ghostine et al. (2021), Wintachai and Prathom (2021), and Antonini, Calandrini, and Bianconi (2021) are age-homogeneous. In the early stages of the pandemic in the UK, Crellen et al. (2021) used an SEIR model to investigate the effect of different mean durations of immunity (90-365 days), on short-term dynamics and long-term endemic behaviour, in an age-based model, in the absence of vaccination, under various assumptions about the time-varying effective reproduction number. In Xu, Wu, and Topcu (2021), the authors consider a short-term SEIR model over 200 days, that optimises vaccination roll-out, subject to thresholds on death rates, cumulative deaths, and target numbers of those vaccinated. It assumes 100% vaccine efficacy, with no waning of immunity following vaccination or infection. Ghostine et al. (2021) developed a SEIR model for short-term predictions in Saudi Arabia. It is age-homogeneous, fits to actual data, and takes account of vaccine efficacy, but not of waning immunity. Wintachai and Prathom (2021) use a SEIR model and study the efficiency of vaccines for Covid-19 situations, again fitting parameters to actual data. Antonini, Calandrini, and Bianconi (2021) use a mean duration of vaccinal and infection immunity of 240 days to predict short-term outcomes in Italy.

Other authors have used heterogeneous SIR/SEIR models. Saad-Roy et al. (2020) develop a Covid-19 SIR based model demonstrating a large range of model outcomes, ranging from elimination to high levels of endemic behaviour, as the speculative biological parameters, including vaccine efficacy and waning rates, are changed. The authors emphasise the importance of characterising these parameters if such models are to be useful for informing management policies. Moore et al. (2021) developed a compartmental model, heterogeneous by age and region, to predict the evolution of Covid-19 under various combinations of vaccine roll-out and time-dependent NPIs in the UK. This pre-dated the delta variant’s emergence as the dominant variant. It does not consider waning of immunity, either from infection or vaccination, or a combination of these two. We try to address these features, which are now recognised to be an important issue, as exemplified by a perceived urgent need to roll out booster doses. A finding of their study is that vaccination alone, even pre-delta, would not contain the virus. Patel et al. (2021) use agent-based simulation within an SEIR framework with heterogeneous transmission and age-specific mortality rates to model decision strategies over an 18-month period in North Carolina. The model does not consider waning immunity. The authors found that removing NPIs while distributing vaccines resulted in a significant increase in the number of infections, hospitalisations, and deaths.

Many of the above are characterised by being short-term predictive studies, prior to emergence of the delta variant, fitting parameters to local data, and with no explicit consideration of waning immunity and vaccine efficacy. It is now generally believed that Covid-19 will be with us for a long time (Scudellari, 2020). At the time of writing, estimates of the efficacy against infection by the delta variant were improving (Pouwels et al., 2021), but there is uncertainty in waning rates and one cannot predict what new more transmissible variants might emerge. In the light of these uncertainties, rather than forecasting, we are interested in understanding what the long-term endemic levels of infection and mortality might be under speculative assumptions about waning immunity, vaccine efficacy, and control reproduction number.

## 3 Model

The model is of a homogeneously mixing population with six compartments representing those who are: susceptible and will not be vaccinated (*S*_1_); susceptible and will be vaccinated (*S*_2_); infected (*I*); vaccinated with no loss of immunity (*V*); recovered (*R*); and died from Covid-19 (*M*), as shown in Figure 1.

**Figure 1:**
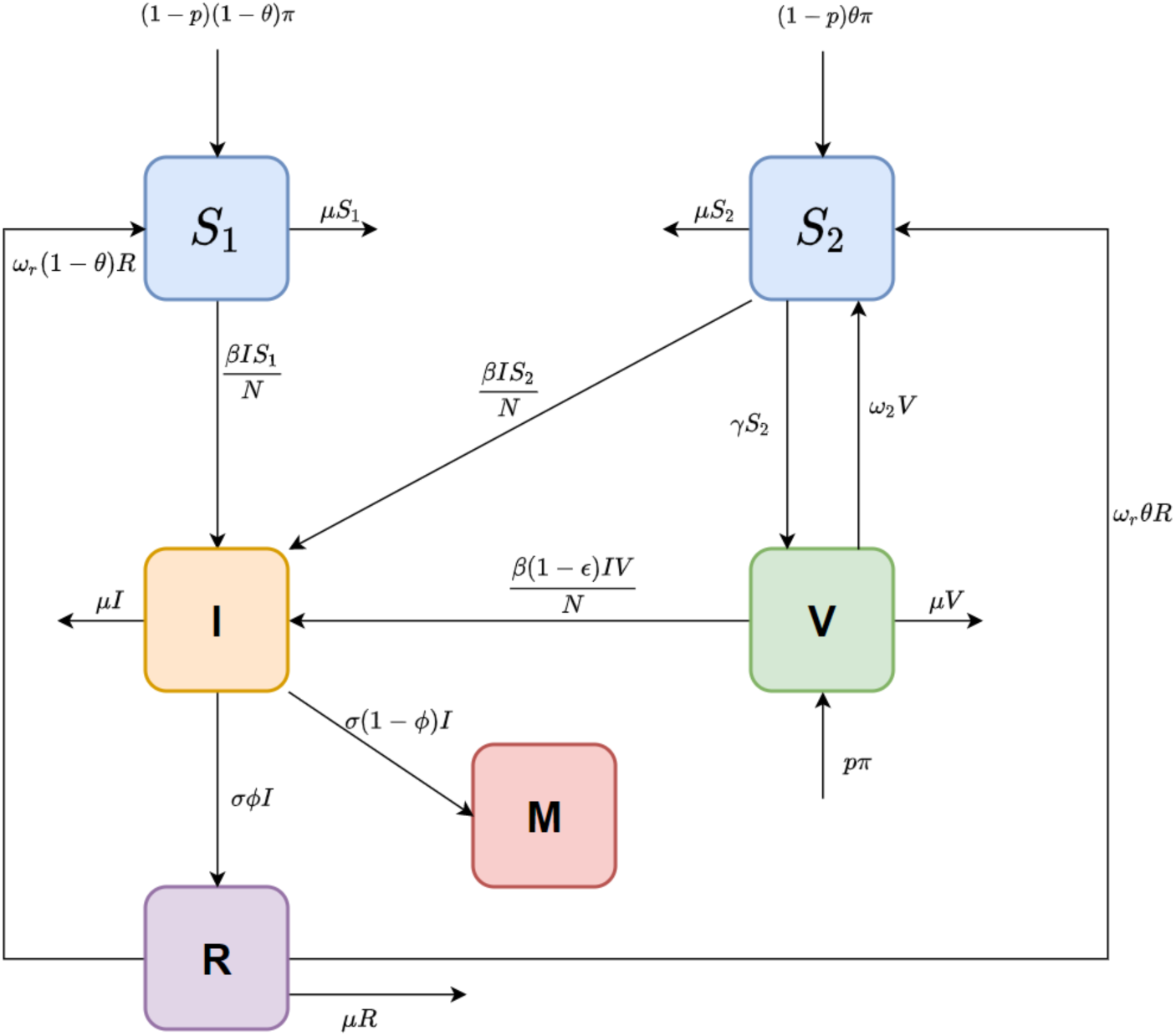
Flow chart of the vaccine model with variables and parameters shown in Table 1.

The model is given by the following ordinary differential equations (ODE):

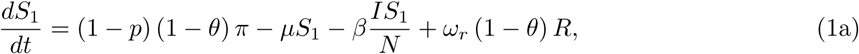

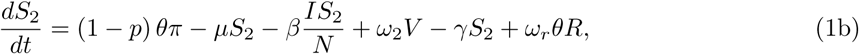

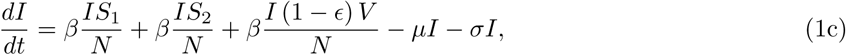

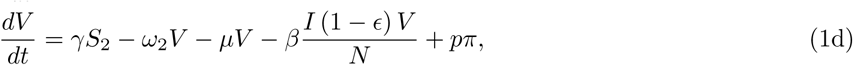

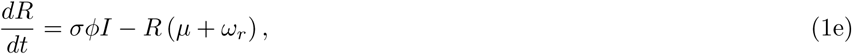

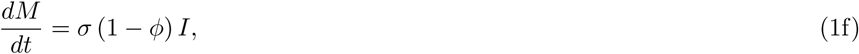

**Table 1:** Description of variables and parameters in the model.

| Variable<br>/Parameter | Interpretation |
| --- | --- |
| $S_1$ | Number of susceptible population who do not aspire to vaccination. |
| $S_2$ | Number of susceptible population who aspire to vaccination. |
| $I$ | Number of population who are infected. |
| $V$ | Number of population who retain immunity after vaccination. |
| $R$ | Number of living population who survived infection. |
| $M$ | Cumulative number of Covid-19 deaths. |
| $N$ | Size of living population ($N = S_1 + S_2 + I + V + R$). |
| $\mu$ | Non-Covid death rate per head of population. |
| $\pi$ | Birth rate of living population. |
| $p$ | $Prob(\text{vaccination at birth})$. |
| $\theta$ | $Prob(\text{a susceptible aspires to vaccination given not vaccinated at birth})$, assumed to be independent of age. |
| $\omega_1$ | Waning rate of immunity against infection alone. |
| $\omega_2$ | Waning rate of immunity against vaccination alone. |
| $\omega_{12}$ | Waning rate of immunity against vaccination and infection. |
| $\omega_r$ | Waning rate of immunity against recovered state. |
| $\gamma$ | Vaccination rate for susceptibles who aspire to vaccination. |
| $\beta$ | Transmission rate. |
| $\sigma$ | Rate of leaving the infected state. |
| $\epsilon$ | Efficacy of vaccine against infection. |
| $1 - \phi$ | $Prob(\text{death given infection})$. |

where

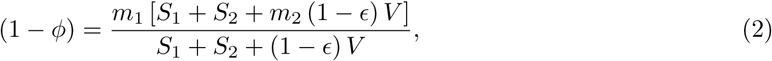

and *m*_1_, *m*_1_*m*_2_ are the infection fatality rates for unvaccinated and vaccinated persons respectively. Similarly,

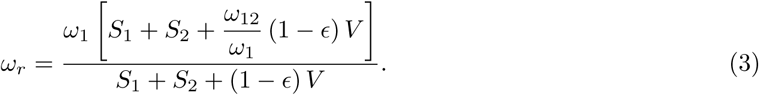

The explanation of variables and parameters in system (1) are presented in Table 1. The system of ODEs (1) are under the conditions that 0 ≤ *S*_1_ (*t*), *S*_2_ (*t*), *I* (*t*), *V* (*t*), *R* (*t*) ≤ *N*.

Adding equations (1a)-(1e) we find that the living population size, *N*, follows

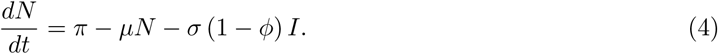

To balance births and deaths, we set

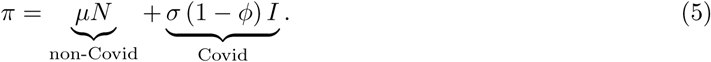

This ensures that the living population is stable over time. The first term on the right-hand side of Eq.(5) arises from non-Covid-19 deaths and the second term from Covid-19 deaths.

The control reproduction number *R*_*c*_ is the average number of secondary infections from a primary infector, under current controls, in a hypothetical completely susceptible population. It is smaller than the basic reproduction number *R*_0_ because of the controls. In seeking to understand long-term evolution of the pandemic, we will assume that *R*_*c*_ does not vary with time. That is, for *t*> 0 the mix of control measures and individual behaviours such as level of homeworking, mask wearing (see for example Chu et al. (2020)), improved ventilation, improved test-trace-isolate methods, are all assumed to result in a constant *R*_*c*_ that is smaller than *R*_0_. This ignores factors such as seasonality and changes in NPIs, such as short-term suppressions and release. The basic reproduction number for the delta variant is perhaps around 6 but we will examine the impact of values of *R*_*c*_ below that, to reflect the new normal of “living with the virus”. We assume, somewhat optimistically, that there will be no new variant that is more transmissible, more virulent, has more vaccine escape than the delta variant. We do not intend the model to be predictive, rather to use it to gain some understanding of the impact of different levels of vaccination coverage, of waning immunity, and of vaccine efficacy, on the long-term infection prevalence and mortality rate of the disease.

For this compartmental model the control reproduction number is

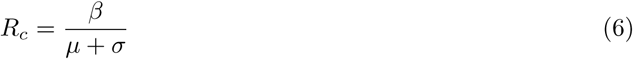

and the effective (or time-dependent) reproduction number is

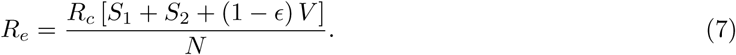

Note from Eq.(1c) and (7) that the prevalence of infections is increasing or decreasing according as *R*_*e*_ < 1, *R*_*e*_ > 1.

A *disease-free equilibrium* 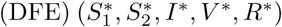 is obtained by setting the derivatives of equations (1a)-(1f) to zero with *I* = 0. Let 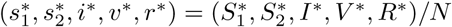 where 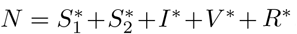. The lower-case state variables now represent the equilibrium proportions of the living population occupying each of the five compartments. The solution is

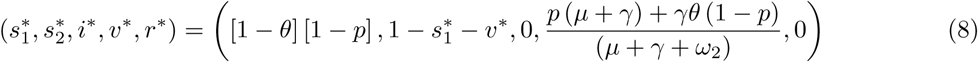

At this DFE solution, 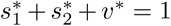 and so from Eq.(7), the effective reproduction number at DFE is

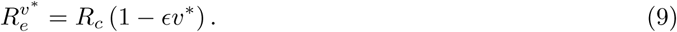

Henceforth, we assume that *µ* = 0, as the non-Covid death rate is small in comparison to waning and vaccination rates. The DFE is stable providing 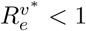, that is providing

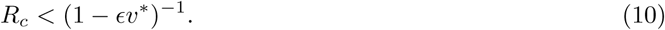

Inequality (10) is the condition for disease elimination in the long run. If it is not satisfied then the disease is endemic and persists for ever. Table 2 shows specimen threshold values of *R*_*c*_ for various parameter values when *p* = 0, corresponding to no vaccination at birth. The chosen values for efficacy (0.65, 0.8) against the delta variant are guided by those given for the Astra Zeneca (Vaxzevria) and Pfizer-BioNTech (Cominarty) vaccines respectively, in the Covid-19 vaccine surveillance report (Public Health England, 2021).

**Table 2:** Thresholds for control reproduction number that enable Disease-Free Equilibrium for *p* = 0 with vaccine efficacy (*ϵ*) against the delta variant to be 0.65 and 0.8.

| Parameter values |  |  | When vaccine efficacy against infection is 0.65 |  | When vaccine efficacy against infection is 0.80 |  |
| --- | --- | --- | --- | --- | --- | --- |
| $\theta$ | $\omega_2^{-1}$ | $\gamma^{-1}$ | $R_c$ threshold | $v^*$ | $R_c$ threshold | $v^*$ |
| 0.65 | 770 | 30 | 1.69 | 0.63 | 2.00 | 0.63 |
| 0.8 | 770 | 30 | 2.00 | 0.77 | 2.60 | 0.77 |
| 0.95 | 770 | 30 | 2.47 | 0.91 | 3.72 | 0.91 |
| 0.65 | 180 | 30 | 1.57 | 0.56 | 1.80 | 0.56 |
| 0.8 | 180 | 30 | 1.80 | 0.69 | 2.22 | 0.69 |
| 0.95 | 180 | 30 | 2.12 | 0.81 | 2.87 | 0.81 |
| 0.65 | 770 | 180 | 1.52 | 0.53 | 1.73 | 0.53 |
| 0.8 | 770 | 180 | 1.73 | 0.65 | 2.08 | 0.65 |
| 0.95 | 770 | 180 | 2.00 | 0.77 | 2.60 | 0.77 |
| 0.65 | 180 | 180 | 1.27 | 0.33 | 1.35 | 0.33 |
| 0.8 | 180 | 180 | 1.35 | 0.40 | 1.47 | 0.40 |
| 0.95 | 180 | 180 | 1.45 | 0.48 | 1.61 | 0.48 |

The threshold is increasing in vaccination coverage, vaccine efficacy, vaccination rate, and decreasing in waning rate. Larger thresholds for *R*_*c*_ are indicative of more easily eliminating the disease, since they become closer but less in value to *R*_0_. We see that the thresholds are considerably smaller than the basic reproduction number for delta, say *R*_0_ = 6. For example, in the first row of table 2, a threshold of *R*_*c*_ = 1.69 might indicate that the NPIs, assuming that is independent of controls, would need to be so strong as to decrease the transmission rate to 28% of what it was, were there no controls. Considering that early in the pandemic in the UK the basic reproduction number was around 3 (Billah, Miah, and Khan, 2020) and brought down to about 0.6 during the first lockdown (20% of the original transmission rate), 28% is achievable, but obviously not in perpetuity, which is the situation we consider here. So, we conclude from table 2 that vaccination alone will certainly not lead to disease elimination of the delta variant. For the wild type of virus (*R*_0_ = 3 say), the threshold of 3.72 in row three of table 2 suggests that elimination might just have been possible by vaccination alone, but only with with very high vaccination coverage, very high efficacy, and very low waning rate. For the case of all vaccinated at birth, *p* = 1, the thresholds turn out to be larger, although it may take longer to reach DFE as the supply rate of new vaccinated persons is small and equal to the Covid death rate, *σi*(1 − *ϕ*).

We now obtain an equilibrium solution 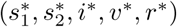 for the endemic case (disease persists in perpetuity) by setting the derivatives in Eq. (1a)-(1e) to zero. Note from (9) that when these derivatives are zero, then 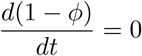 and 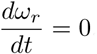.We take *µ* = 0. This leads to

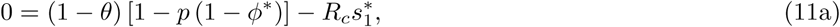

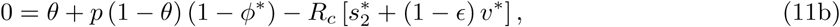

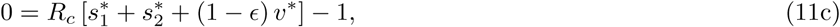

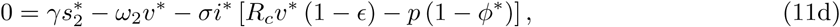

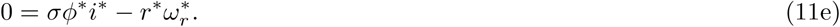

Note from Eq(7) and (11c), that at equilibrium the effective reproduction number *R*_*e*_ = 1. Using Eq(2) and Eq(3), *ϕ*^*^ and 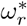 in System (11) can be expressed as

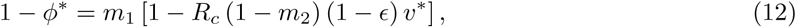

and

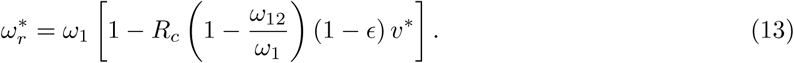

From Eq(1f) the stable endemic death rate, expressed as a proportion of the living population, is

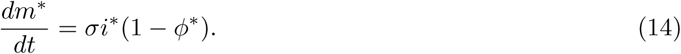

Finally, in the case of no vaccination with *θ* = 0, *p* = 0, the disease will persist if *R*_*c*_ > 1 in which case the stable endemic equilibrium state is

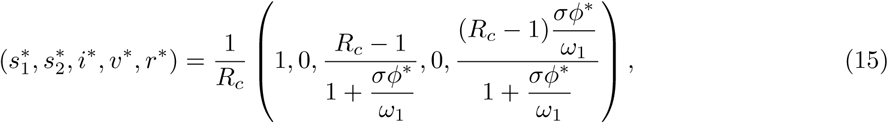

where *ϕ*^*^ = 1 − *m*_1_ with

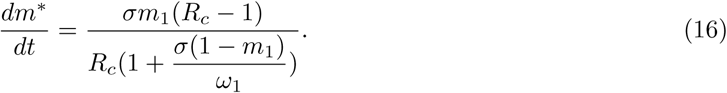

## 4 Results

The model requires input values for parameters, and here we summarise what has been reported on these. In general, estimates of biological parameters have high uncertainty, may not be stationary over time, and can vary with geographical region and age heterogeneity. The recent paper by Pouwels et al. (2021) reported the results of a UK Covid-19 Infection Survey (Office of National Statistics) during a delta-dominant period. Vaccine efficacy against infection for persons aged 18 − 64 for Pfizer-BioNTech (Cominarty) was 85% (79 − 90%) and 75% (70 − 80%) at 14 days and 90 days respectively after second dose vaccination. For Astra Zeneca (Vaxzevria) it was 68% (61 − 73%) and 61% (53 − 68%) respectively. The authors also estimated that for those unvaccinated but previously infected, the implicit efficacy of prior infection was 72% (58 − 82%) which suggests initial protection following infection is not too dissimilar to that after vaccination. Levin et al. (2021) reported that six months after taking the second dose, the immunity against delta variants was substantially decreased, especially in people over 65 years old. Goldberg et al. (2021) examined positive cases in Israel between 11 and 31 July 2021 when delta was dominant, and found that those aged more than 60 were 1.6 times more protected if they had received their second dose in March 2021 compared with those who received it in January 2021. Chemaitelly et al. (2021) found in a Qatar-based study, that Pfizer-BioNTech (Cominarty) efficacy against infection and any variant, decreased substantially from around 77% to 20% between 5 and 7 months after the second dose. In contrast, they found efficacy against severe disease and mortality was maintained at 96% up to six months after the second dose. Bar-On et al. (2021) found that a booster dose of Pfizer-BioNTech (Cominarty) vaccine gave very much decreased chances of infection, hospitalisation and death in Israel in a study of the over 60s.

With respect to vaccine coverage, at the time of writing, around 67% of the UK population is double vaccinated with none in the age group 0-11, and the rate of uptake declining. Vaccination of the elderly and vulnerable was prioritised at the beginning. Coverage remains very much age-dependent; see, for example, Public Health England (2021). Vaccine take-up can be influenced by many factors and hence will change in the future. It varies between countries. According to Dror et al. (2020), “vaccine hesitancy” can relate to concerns and doubts about the safety and efficacy of a vaccine.

Here, we run the model with initial conditions (*s*1(0), *s*2(0), *i*(0), *v*(0), *r*(0)) = (0.15, 0, 0.015, 0.64, 0.195) as a fictitious scenario where 64% and 19.5% of the population have immunity through vaccination and previous infection respectively. Of the remaining 16.5%, 15% are susceptible and unwilling or ineligible for vaccination, while the prevalence of infection is 1.5%. We choose base case values for parameter values and then vary each one in turn, while others remain at their base level. These are *θ* = 0.65, *σ*^*-*1^ = 14 days, *m*_1_ = 0.01, 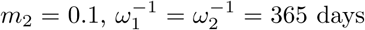, *ω*_12_ = 0.5*ω*_2_, *γ*^*-*1^ = 30 days, *N* = population of UK = 67 million. The vaccine efficacy against infection is *ϵ* = 0.80, against death it is 1 *-m*_2_(1 − *ϵ*) = 0.98, and against death given infection it is 1 − *m*_2_ = 0.90. Estimates of efficacies and waning rates are subject to large uncertainty. The figures used here are guided by the studies referred to previously, by Public Health England (2021) and by Bernal et al. (2021) for the Pfizer-BioNTech (Cominarty) vaccine against the delta variant. We set *ω*_12_ = 0.5*ω*_2_ to introduce some element of prolonged immunity following recovery from infection of a vaccinated person, and then vary that to *ω*_12_ = *ω*_2_. The model does not address age heterogeneity. In the event, in the UK, older people were prioritised in the early stages of vaccine roll-out and so illustrative death rates from this model during the early stages might be expected to be biased to high values.

We set *R*_*c*_ = 4. Such a control reproduction number might be interpreted in one of two ways. Given that *R*_0_ for the delta variant is thought to be around 6, *R*_*c*_ = 4 might be a hypothetical control reproduction number for “living with the virus” in perpetuity. Alternatively, it might be considered as the basic reproduction number for an earlier but less transmissible form of the virus. The model is run over 12 years by which time a stable equilibrium solution to Eq.(1a-1f) was identified for all but one of the examined scenarios.

In figure 2, we show phase diagrams for susceptible proportion and prevalence of infection. In figure 3, the infection prevalence *i*(*t*) and effective reproduction number *R*_*e*_(*t*) are plotted for different scenarios which all lead to stable endemic equilibria, with a limit value of 1 for *R*_*e*_(*t*). In figure 4, the death rate 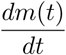 is plotted. In figures 2a and 3a, changing the vaccination coverage *θ* has little effect in the short-run where there is a surge in infections and a large first wave. This would seem to be the result of a large *R*_*c*_, and a reservoir of people, both susceptible and vaccinated, still capable of becoming infected. One might be tempted to avoid that by reducing *R*_*c*_ for small *t*, but that would only delay the inevitable if the intention were to restore it to 4 eventually. In the long-term, the stable endemic levels for *i*(*t*) (figure 3a) and 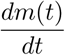 (figure 4a) are very sensitive to the target vaccination coverage (*θ*). Of particular note is the long “honeymoon period” of around 3 years when *θ* = 0.95.

**Figure 2:**
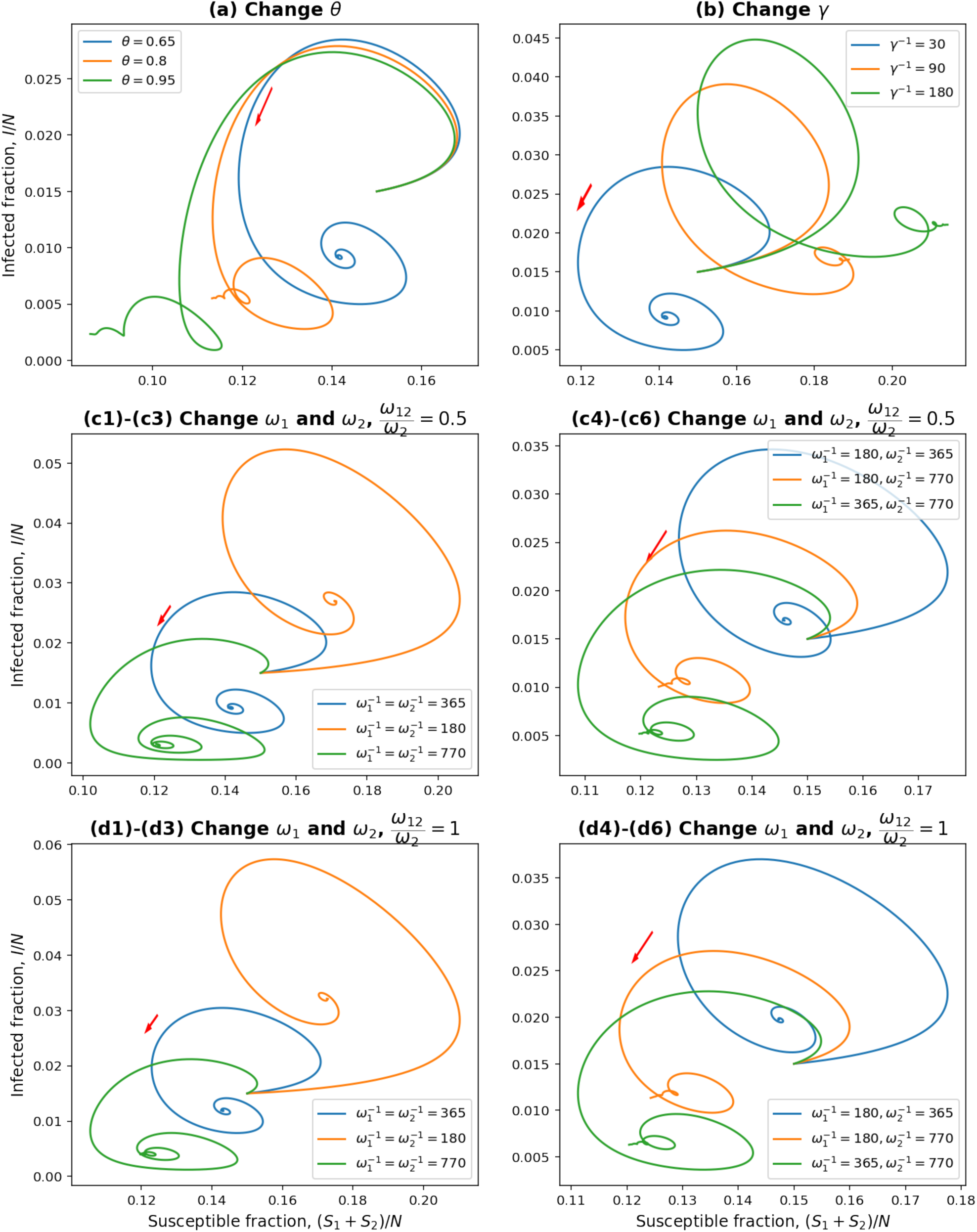
Phase diagrams for different parameters under the *R*_*c*_ = 4 condition.

**Figure 3:**
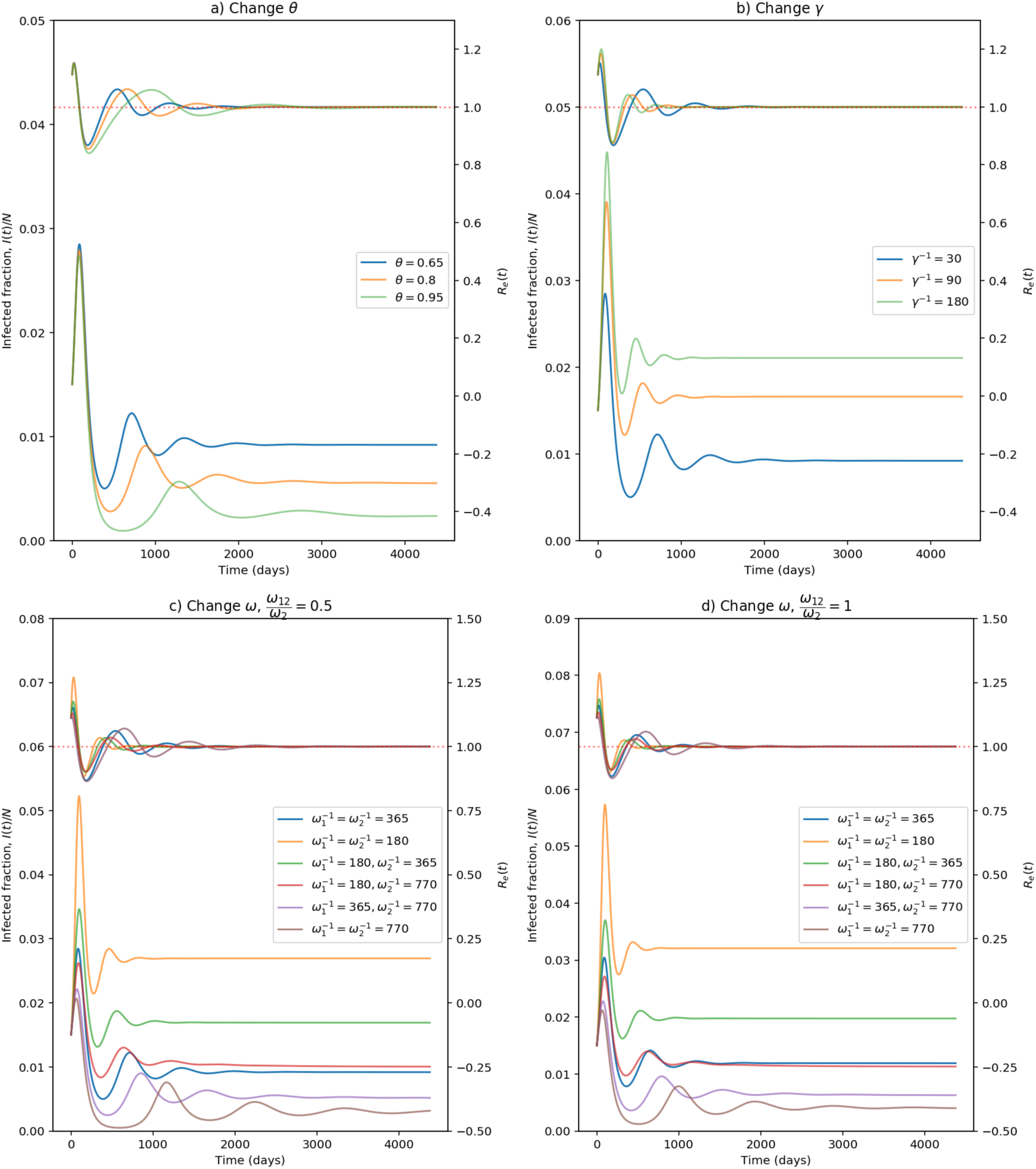
Changes in infection prevalence *i*(*t*) and effective reproduction number *R*_*e*_(*t*) for different parameters under the *R*_*c*_ = 4 condition. The vertical axis for *i*(*t*) shown on the left-hand side and the vertical axis for *R*_*e*_(*t*) shown on the right-hand side. (Note: For c) and d), *ω* means *ω*_1_ and *ω*_2_.)

**Figure 4:**
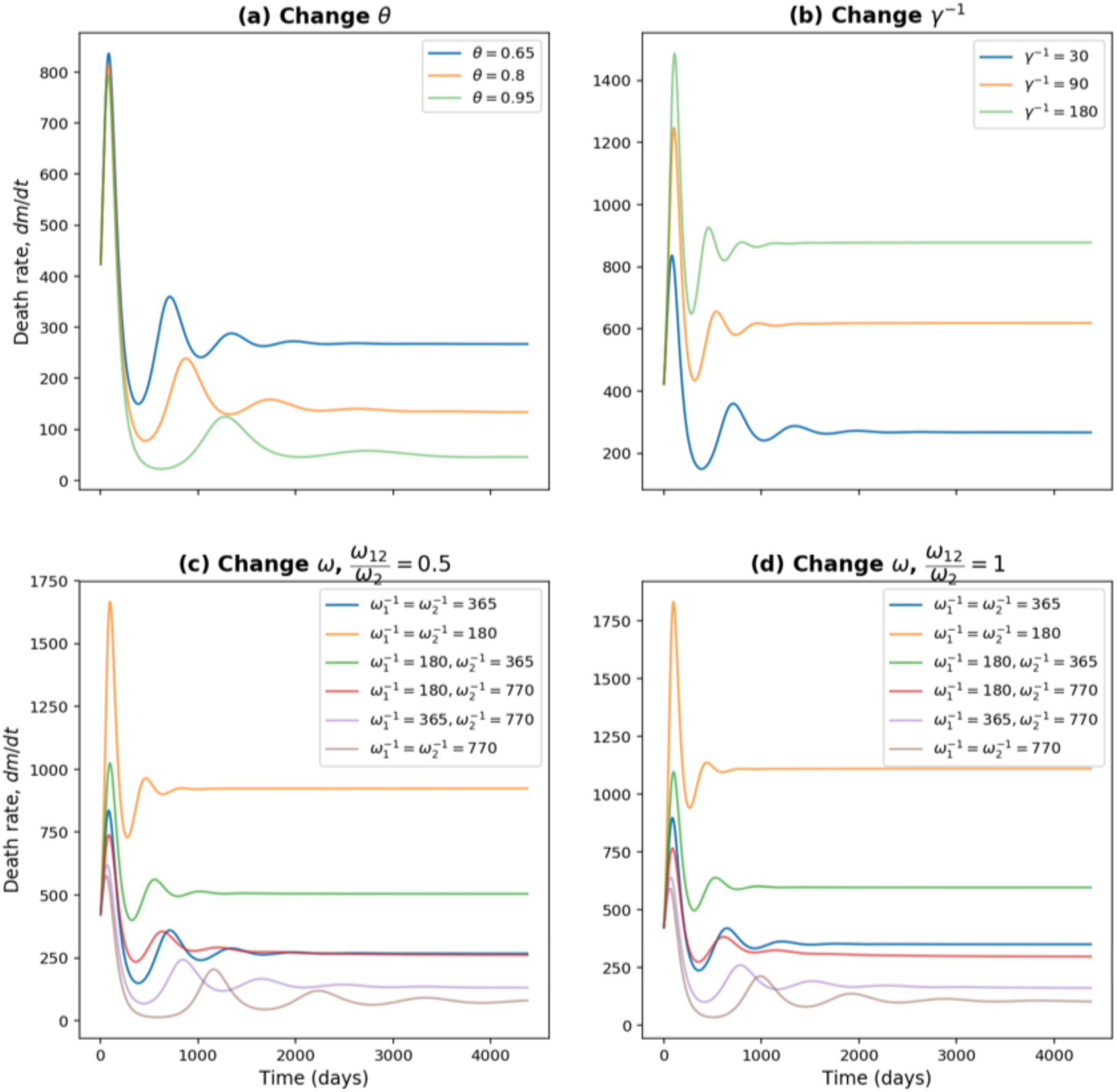
Death rate for different parameters under the *R*_*c*_ = 4 condition. (Note: For c) and d), *ω* means *ω*_1_ and *ω*_2_.)

Increasing the vaccination rate, *γ*, decreases not only the equilibrium prevalence of infected and mortality rates, but also the short-term values. Honeymoon periods increase as *γ* increases and it can take 5 years before near-equilibrium is reached with faster roll-out of vaccines. From figures 3c, 3d, 4c, and 4d, equilibrium levels increase as waning rates increase.When *ϵ* = 0.8, table 3 shows equilibrium mortality rates of between 46 and 1110 per day. In the former case, honeymoon periods are longest and equilibrium is not reached for 10 years. Comparing equilibrium death rates when *ω*_12_ = 0.5*ω*_2_ and *ω*_12_ = *ω*_2_, the extra immunity offered by vaccination followed by infection leads to a reduction of between 12% and 23%.

**Table 3:** A summary of results under different conditions. The last three columns show the day when the peak of infectious cases appears, and numbers in the brackets are the length of honeymoon periods. The basic parameters are: *θ* = 0.65, *ϵ* = 0.8, = *σ* 1*/*14, *m*_1_ = 0.01, *m*_2_ = 0.1, *ω*_1_ = *ω*_2_ = 1*/*365, *γ* = 1*/*30, *R*_*c*_ = 4

| Cases | $R_c$<br>threshold | Experiment results at equilibrium | | | | | | Peak day<br>(length of honeymoon<br>period) | | |
| --- | --- | --- | --- | --- | --- | --- | --- | --- | --- | --- |
| | | $s_1(\infty)$ | $s_2(\infty)$ | $i(\infty)$ | $v(\infty)$ | death<br>rate | death<br>rate<br>without<br>vaccine | 1st | 2nd | 3rd |
| a) Change $\theta$ | | | | | | | | | | |
| a1)<br>0.65 | 1.925 | 0.0876 | 0.0534 | 0.0092 | 0.5453 | 267 | 1339 | 87 | 715<br>(628) | 1348<br>(633) |
| a2)<br>0.8 | 2.447 | 0.0505 | 0.0627 | 0.0055 | 0.6839 | 134 | 1339 | 85 | 882<br>(797) | 1750<br>(868) |
| a3)<br>0.95 | 3.359 | 0.0153 | 0.0708 | 0.0023 | 0.8196 | 46 | 1339 | 83 | 1286<br>(1203) | 2752<br>(1466) |
| b) Change $\frac{1}{\gamma}$ | | | | | | | | | | |
| b1)<br>30 | 1.925 | 0.0876 | 0.0534 | 0.0092 | 0.5453 | 267 | 1339 | 87 | 715<br>(628) | 1348<br>(633) |
| b2)<br>90 | 1.716 | 0.0875 | 0.1014 | 0.0166 | 0.3055 | 619 | 1339 | 104 | 539<br>(435) | 960<br>(421) |
| b3)<br>180 | 1.534 | 0.0875 | 0.1268 | 0.0211 | 0.1786 | 878 | 1339 | 109 | 457<br>(348) | 796<br>(339) |
| c) Change $\frac{1}{\omega}, \frac{\omega_{12}}{\omega_2} = 0.5$ (Shown $\omega_1, \omega_2$ values.) | | | | | | | | | | |
| c1)<br>180,180 | 1.804 | 0.0875 | 0.0838 | 0.0269 | 0.3936 | 924 | 2614 | 98 | 464<br>(366) | 830<br>(366) |
| c2)<br>365,365 | 1.925 | 0.0876 | 0.0534 | 0.0092 | 0.5453 | 267 | 1339 | 87 | 715<br>(628) | 1348<br>(633) |
| c3)<br>770,770 | 2.002 | 0.0918 | 0.0288 | 0.0032 | 0.6498 | 81 | 647 | 63 | 1167<br>(1104) | 2243<br>(1076) |
| c4)<br>180,365 | 1.925 | 0.0875 | 0.0581 | 0.0169 | 0.5222 | 505 | 2614 | 99 | 559<br>(460) | 1023<br>(464) |
| c5)<br>180,770 | 2.002 | 0.0876 | 0.0356 | 0.0101 | 0.6338 | 261 | 2614 | 91 | 641<br>(550) | 1215<br>(574) |
| c6)<br>365,770 | 2.002 | 0.0885 | 0.0312 | 0.0052 | 0.6508 | 132 | 1339 | 73 | 852<br>(779) | 1658<br>(806) |
| d) Change $\frac{1}{\omega}, \frac{\omega_{12}}{\omega_2} = 1$ (Shown $\omega_1, \omega_2$ values.) | | | | | | | | | | |
| d1)<br>180,180 | 1.804 | 0.0875 | 0.0854 | 0.0321 | 0.3854 | 1110 | 2614 | 100 | 441<br>(341) | 781<br>(340) |
| d2)<br>365,365 | 1.925 | 0.0875 | 0.0551 | 0.0119 | 0.5368 | 349 | 1339 | 92 | 651<br>(559) | 1216<br>(565) |
| d3)<br>770,770 | 2.002 | 0.0896 | 0.0301 | 0.0041 | 0.6515 | 102 | 647 | 67 | 999<br>(932) | 1931<br>(932) |
| d4)<br>180,365 | 1.925 | 0.0875 | 0.0597 | 0.0198 | 0.5141 | 596 | 2614 | 103 | 534<br>(431) | 970<br>(436) |
| d5)<br>180,770 | 2.002 | 0.0875 | 0.0367 | 0.0113 | 0.6287 | 297 | 2614 | 94 | 621<br>(527) | 1165<br>(544) |
| d6)<br>365,770 | 2.002 | 0.0881 | 0.0323 | 0.0063 | 0.6480 | 161 | 1339 | 76 | 791<br>(715) | 1533<br>(742) |
| e) Change $\epsilon$. | | | | | | | | | | |
| Continued on next page |  |  |  |  |  |  |  |  |  |  |

Table 3 – continued from previous page
| Cases | $R_c$<br>threshold | Experiment results at equilibrium | | | | | | Peak day<br>(length of honeymoon<br>period) | | |
| --- | --- | --- | --- | --- | --- | --- | --- | --- | --- | --- |
| | | $s_1(\infty)$ | $s_2(\infty)$ | $i(\infty)$ | $v(\infty)$ | death<br>rate | death<br>rate<br>without<br>vaccine | 1st | 2nd | 3rd |
| e1)<br>0.8 | 1.925 | 0.0876 | 0.0534 | 0.0092 | 0.5453 | 267 | 1339 | 87 | 715<br>(628) | 1348<br>(633) |
| e2)<br>0.9 | 2.177 | 0.1044 | 0.0659 | 0.0012 | 0.7878 | 40 | 1336 | 0 | 872<br>(872) | None |
| e3)<br>0.65 | 1.640 | 0.0875 | 0.0434 | 0.0151 | 0.3403 | 413 | 1339 | 70 | 555<br>(485) | 1017<br>(462) |
| e4)<br>0 | 1.00 | 0.2500 | $10^{-18}$ | 0.0280 | $10^{-17}$ | 1339 | 1339 | 30 | 327<br>(297) | 593<br>(266) |
| **f) Change $R_c$.** |  |  |  |  |  |  |  |  |  |  |
| f1) 4 | 1.925 | 0.0876 | 0.0534 | 0.0092 | 0.5453 | 267 | 1339 | 87 | 715<br>(628) | 1348<br>(633) |
| f2) 3 | 1.925 | 0.1261 | 0.0621 | 0.0025 | 0.7240 | 71 | 1188 | 962 | 2072<br>(1110) | None |
| f3) 1.5 | 1.925 | 0.2216 | 0.0591 | $10^{-33}$ | 0.7192 | 0 | 0 | $I(t)$ converges to 0<br>after about 100 days.<br>No honeymoon periods. | | |

In table 3, we show stable equilibrium values of the state variables. *v*^*^ is always always lower than the target coverage *θ*, due to waning immunity and delay time between such loss of immunity and vaccination. The equilibrium death rate is compared with that pertaining to the case of no vaccination. We observe that the percentage reduction is greatest when vaccination coverage is almost universal (*θ* = 0.95) and efficacy is high (*ϵ* = 0.9). In general, it can be seen that with *R*_*c*_ = 4, there are great difficulties in reducing endemic levels, even with a vaccine.

Table 3 shows that the situation would be much easier if *R*_*c*_ were reduced. Results for *R*_*c*_ = 3 appear in figures 5 and 6. Comparing with figures 3 and 4, it is evident that it takes much longer to reach equilibrium and honeymoon periods are larger. From figure 6, it is clear that after 12 years, the death rates are still decreasing for many scenarios we tested here. As expected, equilibrium infection prevalence and mortality rates are much lower. These features are a result of the new *R*_*c*_ being closer to the threshold for disease-free equilibrium, also shown in the table. Meanwhile, from the four plots in figure 5 we find that the *R*_*e*_(*t*) values for *R*_*c*_ = 3 are smaller than those for *R*_*c*_ = 4. In all four plots in figure 3, we can always find the first peak in infection prevalence to be around day 100. But for *R*_*c*_ = 3, we can only find such a feature for the *γ*^*-*1^ = 90 and *γ*^*-*1^ = 180 in figure 5b, 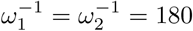, *ω*_12_ = 0.5*ω*_2_ in figure 5c, and 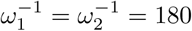, *ω*_12_ = *ω*_2_ in figure 5d. But for other scenarios, the infectious cases experienced a quick decline followed by an increase to a subsequent smaller wave.

**Figure 5:**
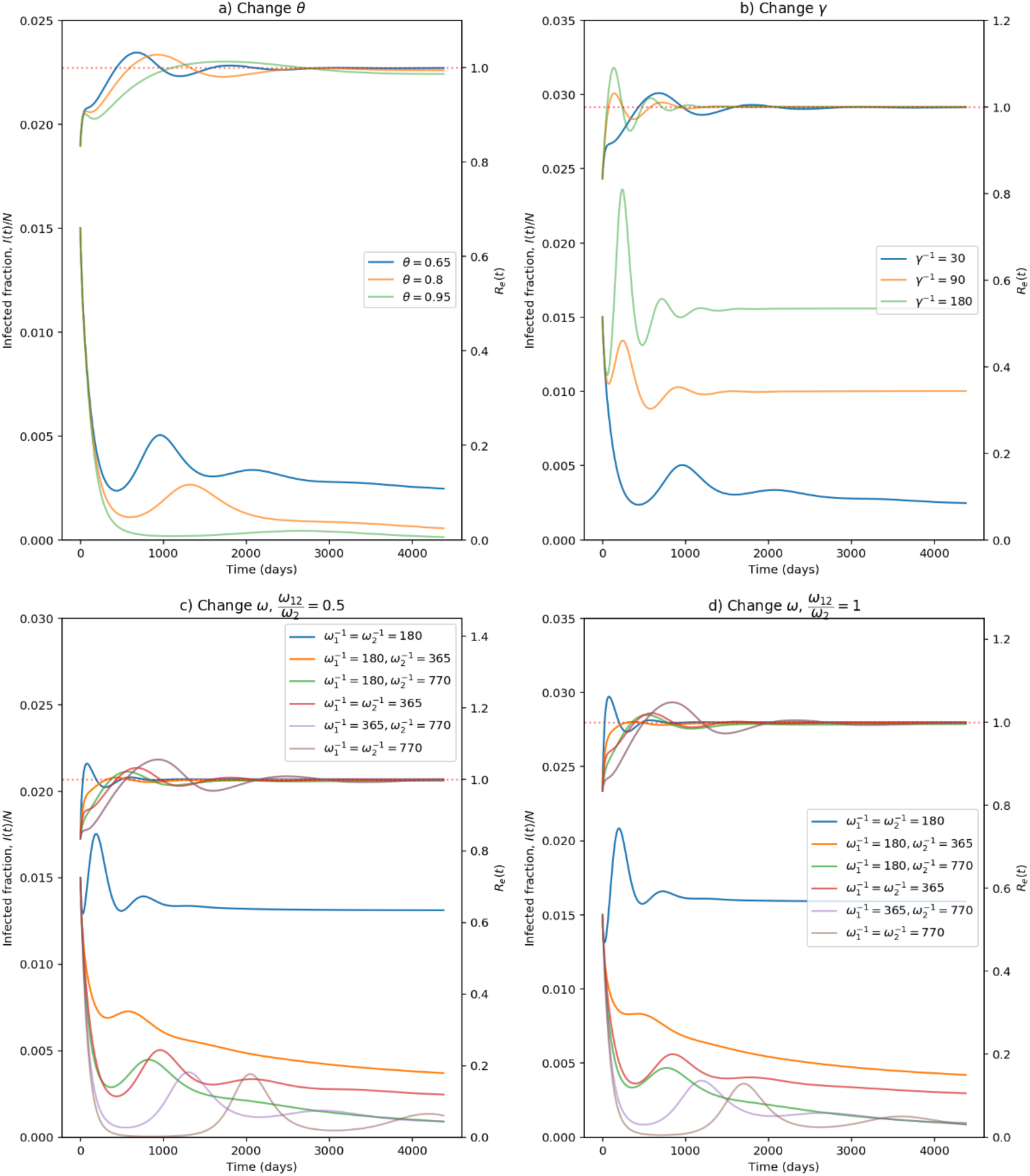
Changes in infection prevalence *i*(*t*) and effective reproduction number *R*_*e*_(*t*) for different parameters under the *R*_*c*_ = 3 condition. The vertical axis for *i*(*t*) shown on the left-hand side and the vertical axis for *R*_*e*_(*t*) shown on the right-hand side. (Note: For c) and d), *ω* means *ω*_1_ and *ω*_2_.)

**Figure 6:**
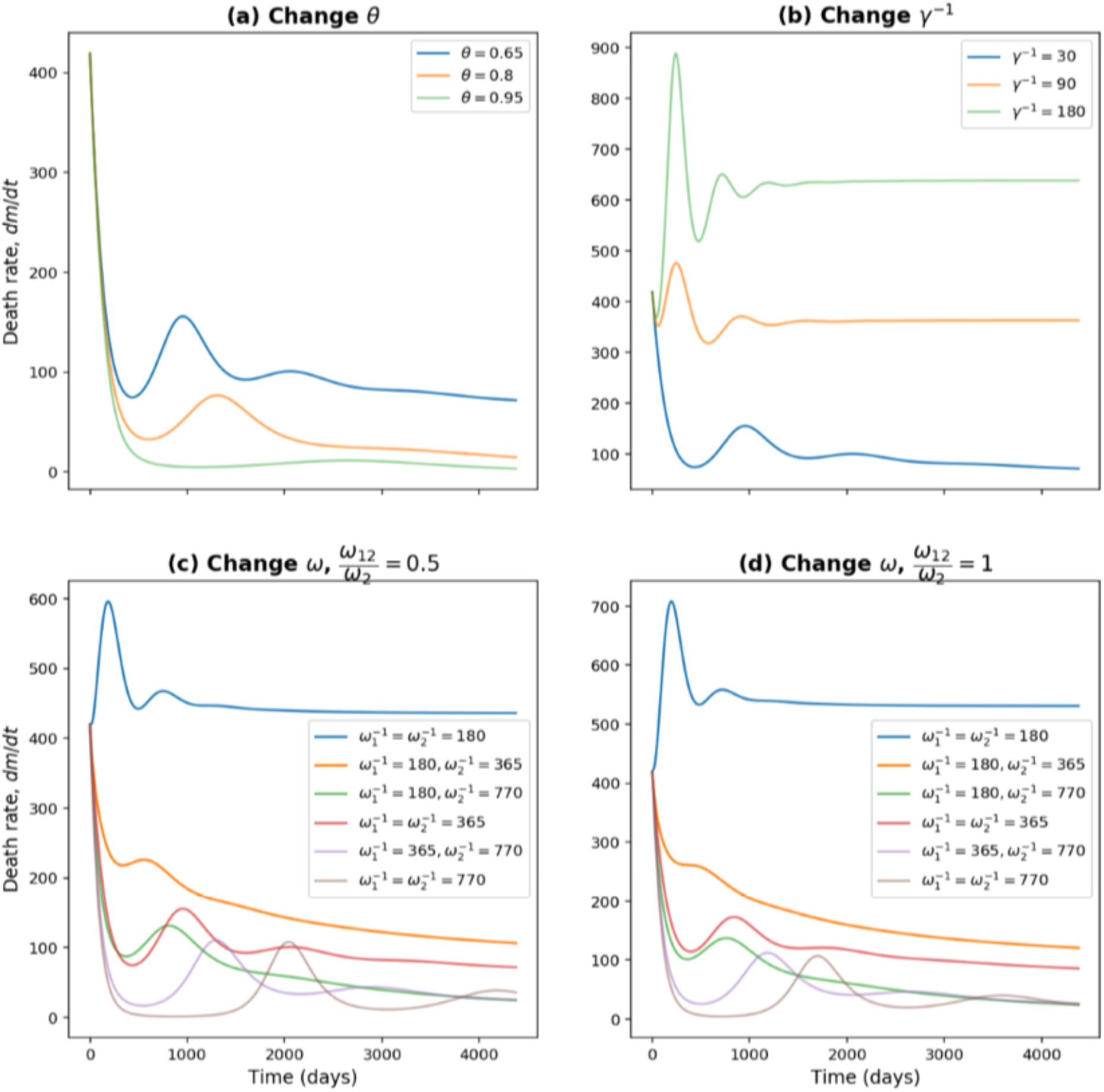
Death rate for different parameters under the *R*_*c*_ = 3 condition. (Note: For c) and d), *ω* means *ω*_1_ and *ω*_2_.)

We have seen that the equilibrium endemic levels can vary hugely depending upon the controllable parameters *θ, γ*, and *R*_*c*_. Vaccines do differ in their efficacy, and in table 3 we show that for the base case of *θ* = 0.65, ^*-*1^ = 14 days, *m*_1_ = 0.01, *m*_2_ = 0.1, 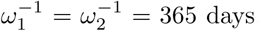, *ω*_12_ = 0.5*ω*_2_, *γ*^*-*1^ = 30 days, the endemic death rate could also vary considerably as (40, 267, 413) per day as the efficacy ranges over (90%, 80%, 65%).

## 5 Discussion and limitations

Results in section 4 illustrate the impact of different parameter values on disease prevalence and death rates. We assumed a control reproduction number of *R*_*c*_ = 4, perhaps in the context of the delta variant with a basic reproduction number of around 6. Comparing the *R*_*c*_ thresholds in table 3 with *R*_*c*_ = 4, highlights the impossibility of attaining disease-free equilibrium. Considering that from Eq. (8) and (10) the condition for DFE is 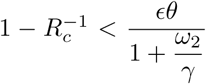, one might aim for a low level of endemic equilibrium by reducing *R*_*c*_ and 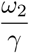 and increasing *ϵ* and *θ*. As we have shown, equilibrium levels are very sensitive to efficacy in the range (0.65, 0.90) and to mean waning periods in the range (180, 770) days, and so this does mean that vaccine design can impact greatly. Similarly, it is important to quickly vaccinate those who are newly susceptible, and our results show that might alter equilibrium levels by a factor of around 3. The best choice would be *γ* = 1*/*30 but that would be challenging. For example, with *s*_2_(*∞*) = 0.053 in the first row of table 3 that would require vaccinations in the UK at the rate of 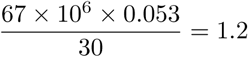 million per day. Increasing *θ* by reducing vaccination hesitancy and extending vaccination to all age groups is important. From the first three rows of table 3, as *θ* increases from 0.65 to 0.95, equilibrium levels change by a factor of around 5. Finally, the major reduction in levels achieved by reducing *R*_*c*_ from 4 to 3, emphasises that non-pharmaceutical interventions (NPIs) should continue to be as important as vaccines in the control toolbox.

A surprising result is that in equilibrium, while an individual’s odds ratio (vaccination versus no vaccination) for death given infection is taken as *m*_2_ = 0.1, the corresponding odds ratio for the population as a whole in equilibrium, is nowhere near that. From Eq. (12), it is 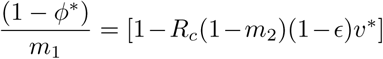. Taking the first three rows of table 3 as an example, the population odds ratios are 0.41, 0.51, 0.61 for *θ* = 0.95, 0.8, 0.65 respectively. These are larger than one might hope for, pointing to the importance of maintaining high values of (1 − *ϵ*)*v*^*^. It seems that although a vaccine might have efficacy against death for an individual of 1 − *m*_2_(1− *ϵ*) = 1 − 0.1(1 − 0.8) = 0.98, much of the reduction in equilibrium death rates across the population comes from the reduction in equilibrium prevalence of infectives and a limited amount from *m*_2_, the inefficacy against death given infection.

A limitation of our model is that it is age-homogeneous. For example, we assume that *θ*, the target vaccination coverage, is the same for all ages, whereas in fact it has been larger for elderly and vulnerable people who have higher infection fatality rates. This would bias death rates to high values. In practice, one might eventually hope for a uniformly high rate of *θ* throughout the age and vulnerability spectrum. Meanwhile, waning rates (*ω*) and vaccination rate (*γ*) in the real world are likely to be age-dependent. Additionally, we have assumed that transmissibility is the same for vaccinated and unvaccinated infectives. To overcome such limitations, one might develop an agent-based simulation model which could also include a more nuanced treatment of continuous waning of immunity, rather than the implicit binary waning of our compartmental model. One might expect such an approach to yield different results to ours in the short-term, but it remains to be seen whether the same is true in estimating equilibrium behaviour.

## 6 Conclusion

In conclusion, three main findings can be drawn from our study. Firstly, the key finding is that vaccination alone would not eliminate the disease. Assuming a control reproduction number of 4, there would eventually be stable endemic levels of disease prevalence and mortality rate. These levels would only be attained after lengthy periods, extending in some cases to 10 years, with a long honeymoon period of between 1 and 3 years between first and second waves. Subsequent waves would have smaller peaks, and in most cases, after the third such wave, levels would be close to equilibrium values. These endemic levels vary widely according to assumed parameter values. For example, with a 95% target vaccination coverage and fast vaccination roll-out, the equilibrium death rate might be 46 per day, compared with 1339 under no vaccination. At the other extreme, with 65% target vaccination coverage and slow vaccination rate, equilibrium death rates might be 878 per day. The equilibrium death rates are very sensitive to the waning : vaccination rate ratio (i.e. *ω* : *γ*). In particular, death rates under low waning might be less than 10% of those with high waning rates. Therefore, good estimates of waning rate will in future be vital to inform policy on suitable control reproduction numbers. Meanwhile, for new Covid-19 vaccine development, high efficacy and low waning rates are obvious and important goals.

Secondly, a chosen control reproduction number is likely to be larger than the threshold value for DFE/Endemic behaviour. The closer it is to the threshold, the lower will be the equilibrium prevalence of infection. But, attaining that equilibrium can take a long time with long honeymoon periods of low prevalence which might be mistaken for disease-free equilibrium.

Thirdly, it is noticeable that the equilibrium values of *v*^*^ are always below the target vaccination rate *θ*. This is an inevitable consequence of waning immunity and time required to vaccinate those who are newly susceptible.

When the Covid-19 vaccination programme was first implemented, perhaps many people thought it would end the pandemic. However, from both our findings and the real data we have in the UK now, it is clear that this is not so. Under a current situation with a highly infectious delta variant, control policies including NPIs are still indispensable to reduce the effective reproduction number. The current policy of allowing high disease prevalence in the UK increases the chance of new more transmissible variants, and these model results demonstrate how the subsequent control becomes more difficult as a result.

## Data Availability

All data produced in the present work are contained in the manuscript

